# The infection fatality rate of COVID-19 inferred from seroprevalence data

**DOI:** 10.1101/2020.05.13.20101253

**Authors:** John P.A. Ioannidis

## Abstract

**Objective:** To estimate the infection fatality rate of coronavirus disease 2019 (COVID-19) from data of seroprevalence studies.

**Methods:** Population studies with sample size of at least 500 and published as peer-reviewed papers or preprints as of July 11, 2020 were retrieved from PubMed, preprint servers, and communications with experts. Studies on blood donors were included, but studies on healthcare workers were excluded. The studies were assessed for design features and seroprevalence estimates. Infection fatality rate was estimated from each study dividing the number of COVID-19 deaths at a relevant time point by the number of estimated people infected in each relevant region. Correction was also attempted accounting for the types of antibodies assessed. Secondarily, results from national studies were also examined from preliminary press releases and reports whenever a country had no other data presented in full papers of preprints.

**Results:** 36 studies (43 estimates) were identified with usable data to enter into calculations and another 7 preliminary national estimates were also considered for a total of 50 estimates. Seroprevalence estimates ranged from 0.222% to 47%. Infection fatality rates ranged from 0.00% to 1.63% and corrected values ranged from 0.00% to 1.31%. Across 32 different locations, the median infection fatality rate was 0.27% (corrected 0.24%). Most studies were done in pandemic epicenters with high death tolls. Median corrected IFR was 0.10% in locations with COVID-19 population mortality rate less than the global average (<73 deaths per million as of July 12, 2020), 0.27% in locations with 73-500 COVID-19 deaths per million, and 0.90% in locations exceeding 500 COVID-19 deaths per million. Among people <70 years old, infection fatality rates ranged from 0.00% to 0.57% with median of 0.05% across the different locations (corrected median of 0.04%).

**Conclusions:** The infection fatality rate of COVID-19 can vary substantially across different locations and this may reflect differences in population age structure and case-mix of infected and deceased patients as well as multiple other factors. Estimates of infection fatality rates inferred from seroprevalence studies tend to be much lower than original speculations made in the early days of the pandemic.

---

The infection fatality rate (IFR), the probability of dying for a person who is infected, is one of the most critical and most contested features of the coronavirus disease 2019 (COVID-19) pandemic. The expected total mortality burden of COVID-19 is directly related to the IFR. Moreover, justification for various non-pharmacological public health interventions depends crucially on the IFR. Some aggressive interventions that potentially induce also more pronounced collateral harms^1^ may be considered appropriate, if IFR is high. Conversely, the same measures may fall short of acceptable risk-benefit thresholds, if the IFR is low.

Early data from China, adopted also by the World Health Organization (WHO),^2^ focused on a crude case fatality rate (CFR) of 3.4%; CFR is the ratio of COVID-19 deaths divided by the number of documented cases, i.e. patients with symptoms who were tested and found to be PCR-positive for the virus. The WHO envoy who visited China also conveyed the message that there are hardly any asymptomatic infections.^3^ With a dearth of asymptomatic infections, the CFR approximates the IFR. Other mathematical models suggested that 40-70%,^4^ or even^5^ 81% of the global population would be infected. Influential mathematical models^5,6^ eventually dialed back to an IFR of 1.0% or 0.9%, and these numbers long continued to be widely cited and used in both public and scientific circles. The most influential of these models, constructed by Imperial College estimated 2.2 million deaths in the USA and over half a million deaths in the UK in the absence of lockdown measures.^5^ Such grave predictions justifiably led to lockdown measures adopted in many countries. With 0.9% assumed infection fatality rate and 81% assumed proportion of people infected, the prediction would correspond to a global number of deaths comparable with the 1918 influenza, in the range of 50 million fatalities.

Since late March 2020, many studies have tried to estimate the extend of spread of the virus in various locations by evaluating the seroprevalence, i.e. how many people in population samples have developed antibodies for the virus. These studies can be useful because they may inform about the extend of under-ascertainment of documenting the infection based on PCR testing. Moreover, they can help obtain estimates about the IFR, since one can divide the number of observed deaths by the estimated number of people who are inferred to have been infected.

At the same time, seroprevalence studies may have several caveats in their design, conduct, and analysis that may affect their results and their interpretation. Here, data available as of July 11, 2020 were collected, scrutinized, and used to infer estimates of IFR in different locations where these studies have been conducted.

## METHODS

### Seroprevalence studies

The input data for the calculations of IFR presented here are studies of seroprevalence of COVID-19 that have been done in the general population, or in samples that might approximate the general population (e.g. with proper reweighting) and that have been published in peer-reviewed journals or have been submitted as preprints as of July 11, 2020. Only studies with at least 500 assessed samples were considered, since smaller datasets would entail extremely large uncertainty for any calculations to be based on them. When studies focused on making seroprevalence assessments at different time interval, they were eligible if at least one time interval assessment had a sample size of at least 500 participants; among different eligible time points, the one with the highest seroprevalence was selected, since seroprevalence may decrease over time as antibody titers wane. Studies with data collected over more than a month, and that could not be broken into at least one eligible time interval that did not exceed one month in duration were excluded, since it would not be possible to estimate a point seroprevalence with any reliability. Studies were eligible regardless of the exact age range of included participants, but studies including only children were excluded.

Studies where results were only released through press releases were not considered here, since it is very difficult to tell much about their design and analysis, and this is fundamental in making any inferences based on their results. Nevertheless, secondarily, results from national studies were also examined from preliminary press releases and reports whenever a country had no other data presented in full papers of preprints as of July 11, 2020. This allowed these countries to be represented in the collected data, but extra caution is required given the preliminary nature of this information. Preprints should also be seen with caution since they have not been yet fully peer-reviewed (although some of them have already been revised based on very extensive comments from the scientific community). However, in contrast to press releases, preprints typically offer at least a fairly complete paper with information about design and analysis.

Studies done of blood donors were eligible, although it is possible they may underestimate seroprevalence and overestimate IFR due to healthy volunteer effect. Studies done on health care workers were not eligible, since they deal with a group at potentially high exposure risk which may lead to seroprevalence estimates much higher than the general population and thus implausibly low IFR. For a similar reason, studies focused on communities (e.g. shelters or religious or other shared-living communities) were also excluded. Studies were eligible regardless of whether they aimed to evaluate seroprevalence in large or small regions, provided that the population of reference in the region was at least 5000 people.

Searches were made in PubMed (LitCOVID), medRxiv, bioRxiv, and Research Square using the terms “seroprevalence” and “antibodies” with continuous updates (last update July 11, 2020). Communication with colleagues who are field experts sought to ascertain if any major studies might have been missed.

Information was extracted from each study on location, recruitment and sampling strategy, dates of sample collection, sample size, types of antibody used (IgG, IgM, IgA), estimated crude seroprevalence (positive samples divided by all samples test), and adjusted seroprevalence and features that were considered in the adjustment (sampling process, test performance, presence of symptoms, other).

### Calculation of inferred IFR

Information on the population of the relevant location was collected from the papers. Whenever it was missing, it was derived based on recent census data trying to approximate as much as possible the relevant catchment area (e.g. region(s) or county(ies)), whenever the study did not pertain to an entire country. Some studies targeted specific age groups (e.g. excluding elderly people and/or excluding children) and some of them made inferences on number of people infected in the population based on specific age groups. For consistency, the entire population, as well as, separately, only the population with age <70 years were used for estimating the number of infected people. It was assumed that the seroprevalence would be similar in different age groups, but significant differences in seroprevalence according to age strata that had been noted by the original authors were also recorded to examine the validity of this assumption.

The number of infected people was calculated multiplying the relevant population with the adjusted estimate of seroprevalence. Whenever an adjusted seroprevalence estimate had not been obtained, the unadjusted seroprevalence was used instead. When seroprevalence estimates with different adjustments were available, the analysis with maximal adjustment was selected.

For the number of COVID-19 deaths, the number of deaths recorded at the time chosen by the authors of each study was selected, whenever the authors used such a death count up to a specific date to make inferences themselves. If the choice of date had not been done by the authors, the number of deaths accumulated until after 1 week of the mid-point of the study period was chosen. This accounts for the differential delay in developing antibodies versus dying from the infection. It should be acknowledged that this is an averaging approximation, because some patients may die very soon (within <3 weeks) after infection (and thus are overcounted), and others may die very late (and thus are undercounted due to right censoring).

The inferred IFR was obtained by dividing the number of deaths by the number of infected people for the entire population, and separately for people <70 years old. The proportion of COVID-19 deaths that occurred in people <70 years old was retrieved from situational reports for the respective countries, regions, or counties in searches done in June 3-7 for studies published until June 7 and in July 3-11 for studies published later. A corrected IFR is also presented, trying to account for the fact that only one or two types of antibodies (among IgG, IgM, IgA) might have been used. Correcting seroprevalence upwards (and inferred IFR downwards) by 1.1-fold for not performing IgM measurements and similarly for not performing IgA measurements may be reasonable, based on some early evidence,^7^ although there is uncertainty about the exact correction factor.

### Data synthesis considerations

Inspection of the IFR estimates across all locations showed vast heterogeneity with heterogeneity I^2^ exceeding 99.9% and thus a meta-analysis would be inappropriate to report across all locations. Quantitative synthesis with meta-analysis across all locations would also be misleading since locations with high seroprevalence would tend to carry more weight than locations with low seroprevalence; locations with more studies (typically those that have attracted more attention because of high death tolls and thus high IFRs) would be represented multiple times in the calculations; and more sloppy studies with fewer adjustments would get more weight, because they would have spuriously tighter confidence intervals than more rigorous studies with more careful adjustments allowing for more uncertainty. Finally, with a highly skewed IFR distribution and with extreme between-study heterogeneity, synthesis with a typical random effects model would tend to produce an erroneously high summary IFR that approximates the mean of the study-specific estimates (also heavily driven by hotbed high-mortality locations with more studies done), while for a skewed distribution the median is more appropriate.

Therefore, at a first step, IFR estimates from studies done in the same country (or in the US, the same state) were grouped together and a single IFR was obtained for that location, weighting the study-specific IFRs by the sample size of each study. This allowed to avoid giving inappropriately more weight to studies with higher seroprevalence estimates and those with seemingly tighter confidence intervals because of poor or no adjustments, while still giving more weight to larger studies. Then, a single summary estimate was used for each location and the median of the distribution of location-specific IFR estimates was calculated. Finally, it was explored whether the location-specific IFRs were associated with the COVID-19 mortality rate in the population (COVID-19 deaths per million people) in each location as of July 12, 2020; this allowed to assess whether IFR estimates tend to be higher in harder hit locations.

## RESULTS

### Seroprevalence studies

36 studies with a total of 43 eligible estimates were published either in the peer-reviewed literature or as preprints as of July 11, 2020.^8-43^ Dates and processes of sampling and recruitment are summarized in Table 1, sample sizes, antibody types assessed and regional population appear in Table 2, estimated prevalence, and number of people infected in the study region are summarized in Table 3, and number of COVID-19 and inferred IFR estimates are found in Table 4. Several studies performed repeated seroprevalence surveys at different time points, and only the time point with the highest seroprevalence estimate is considered in the calculations. With three exceptions, this is also the latest time point. Furthermore, another 7 preliminary national estimates were also considered (Table 5)^44-50^ from countries that had no other seroprevalence study published as a full paper or preprint. This yielded a total of 50 eligible estimates.

**Table 1.**
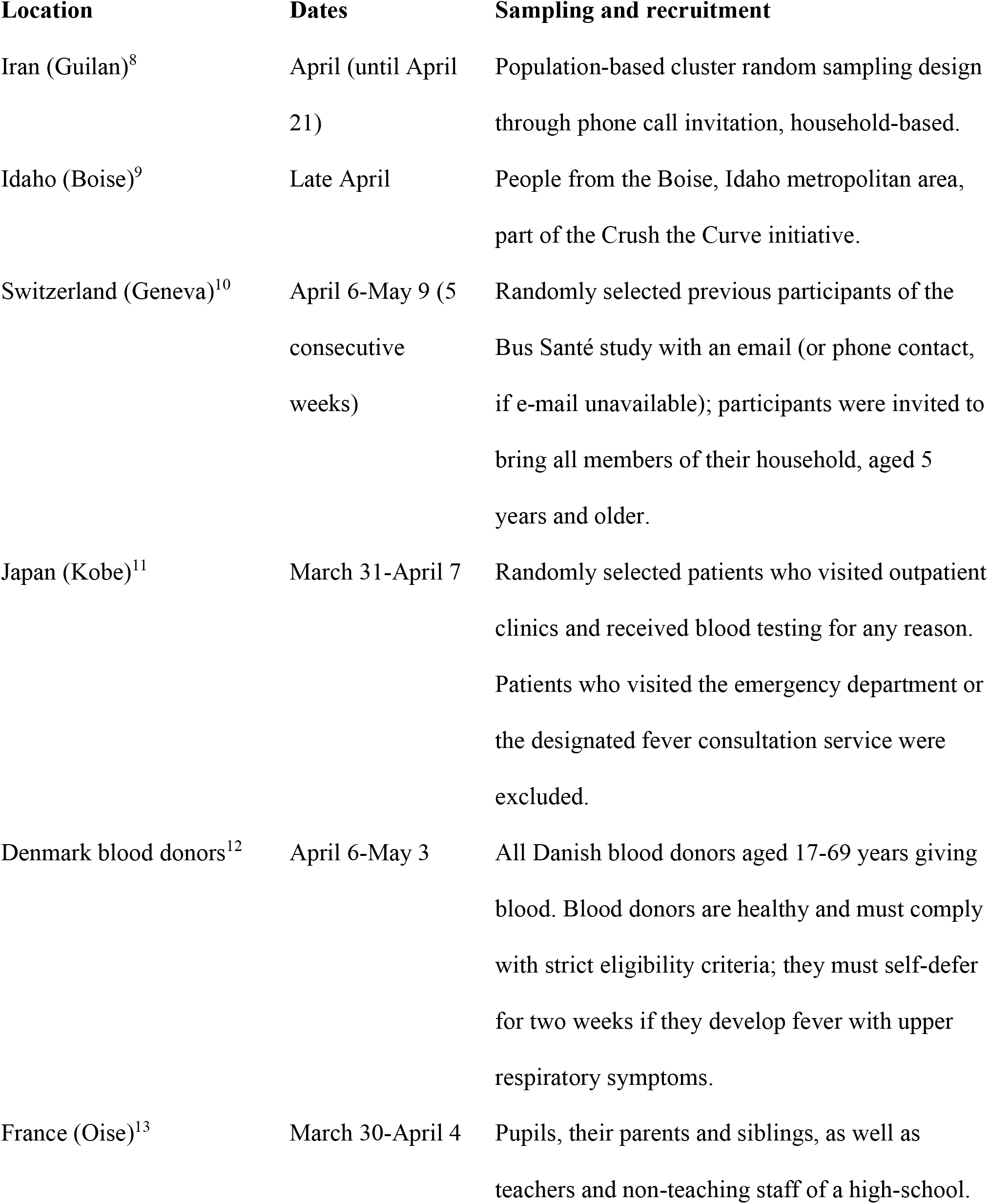
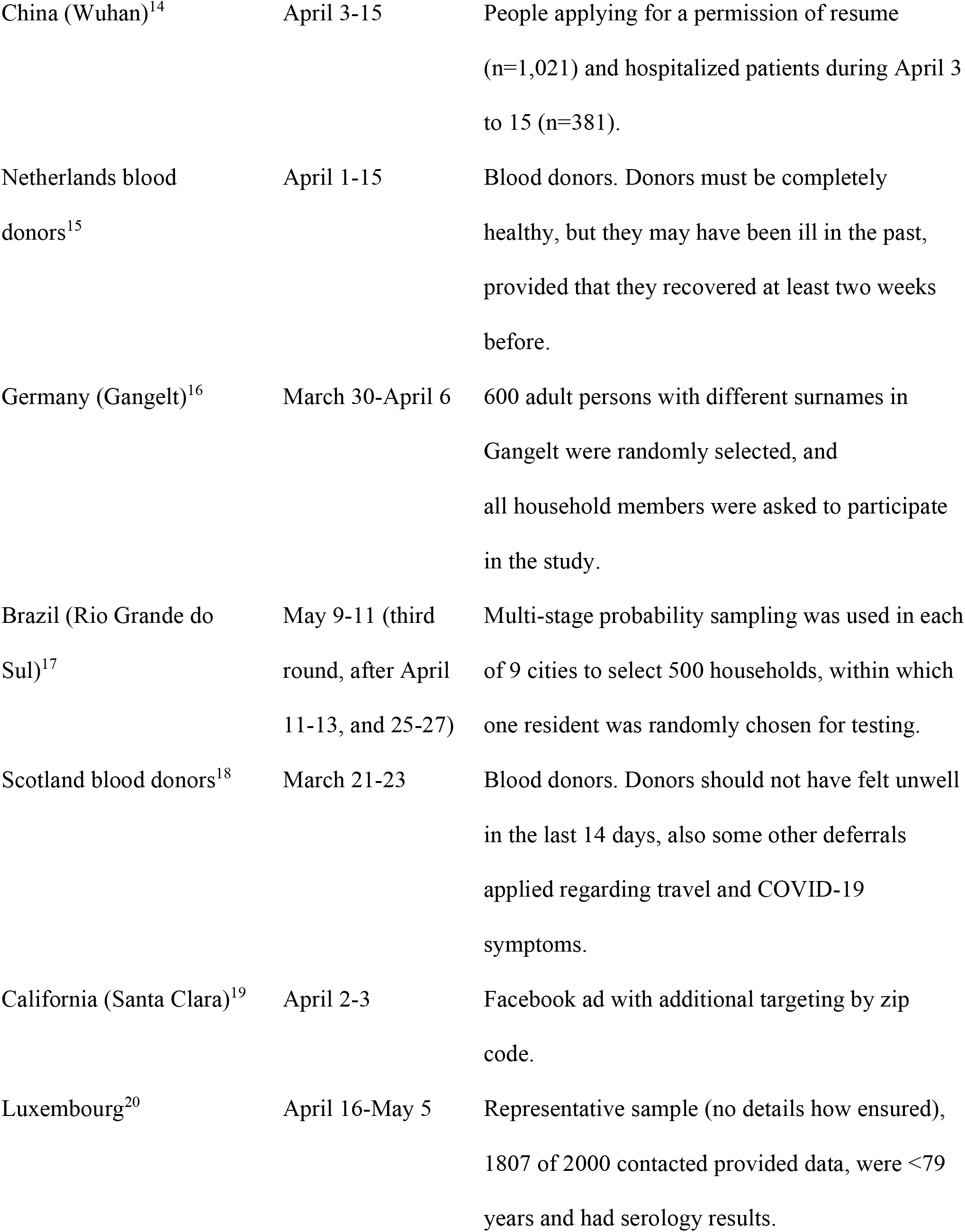
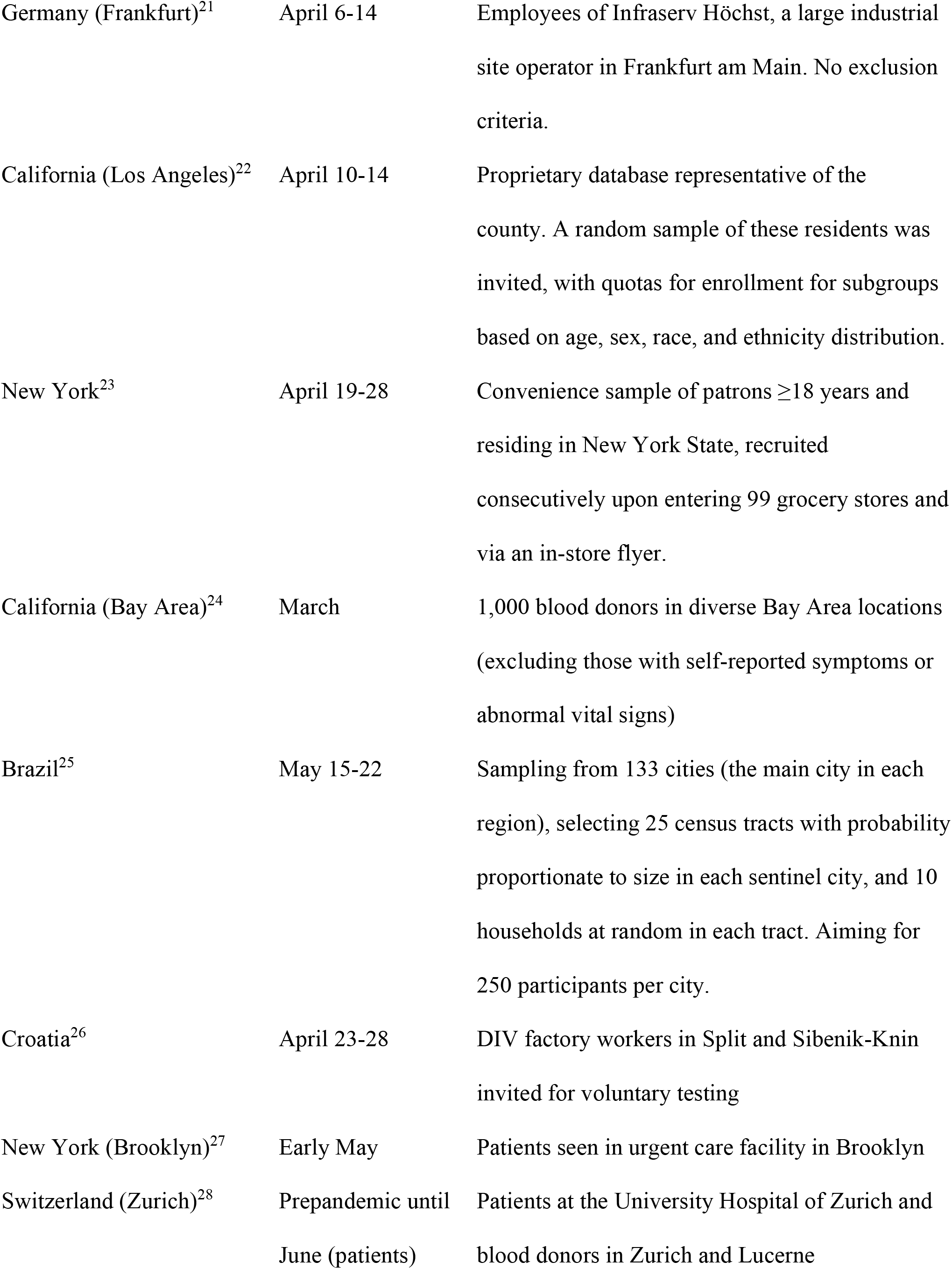
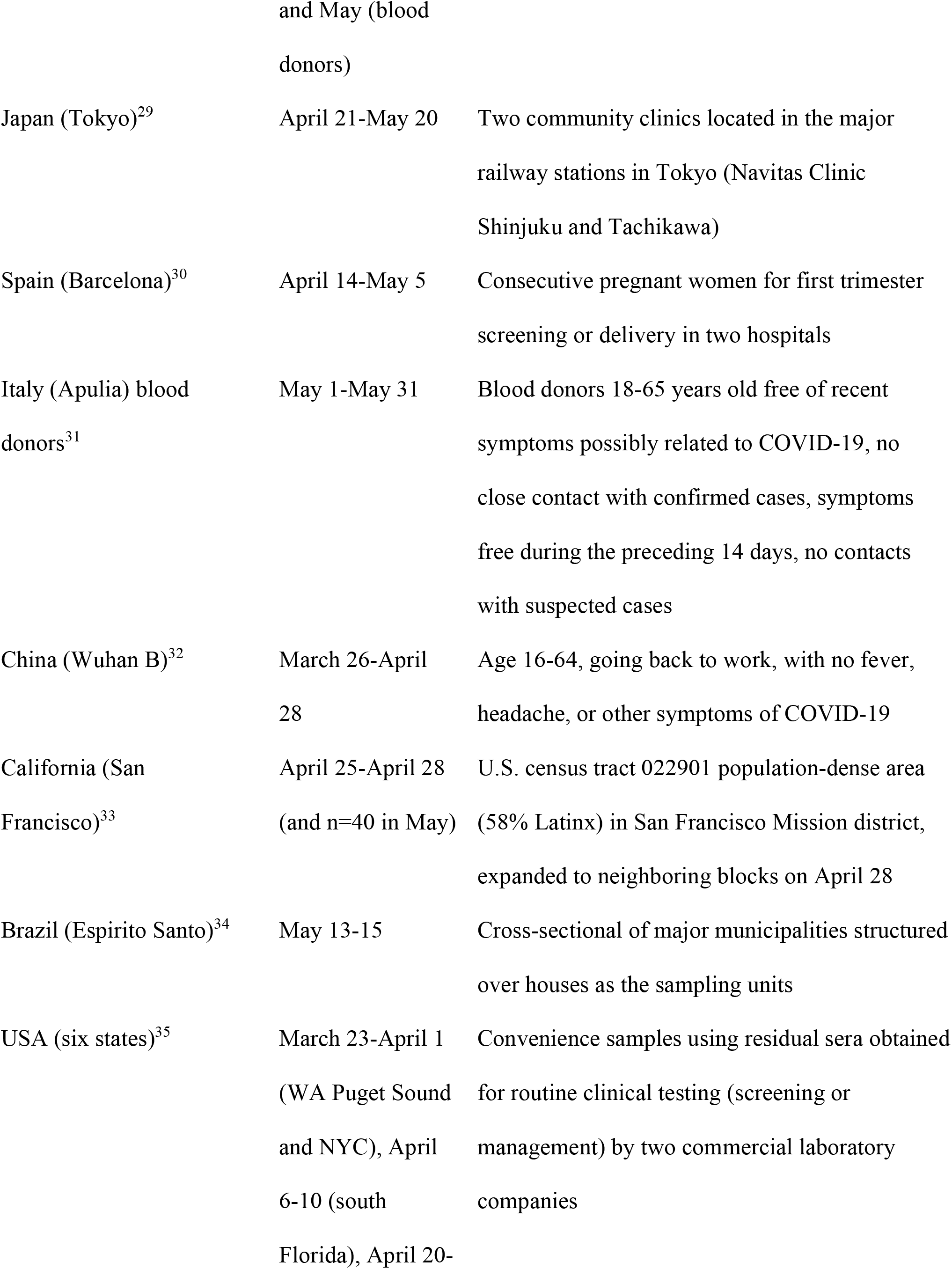
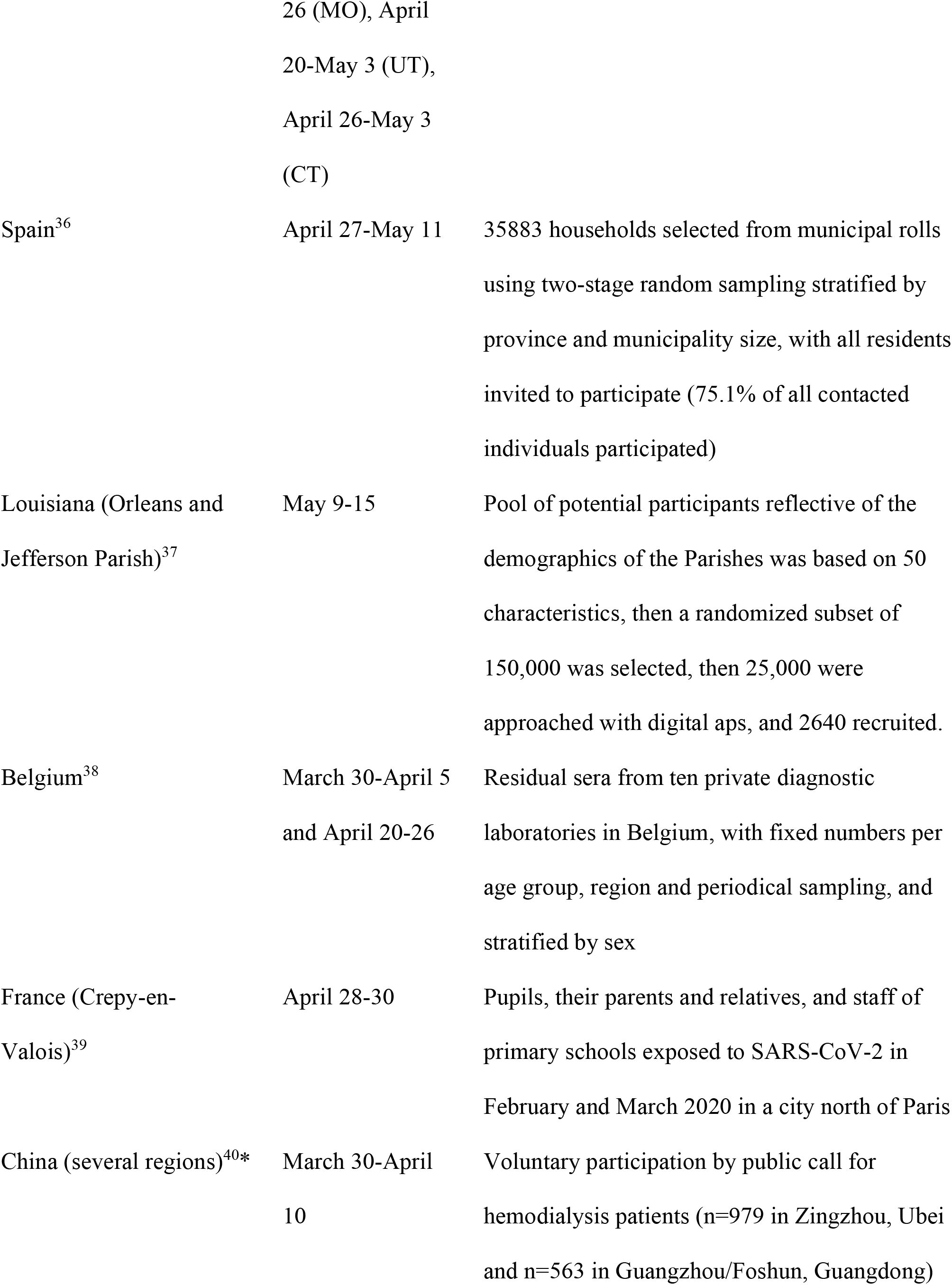
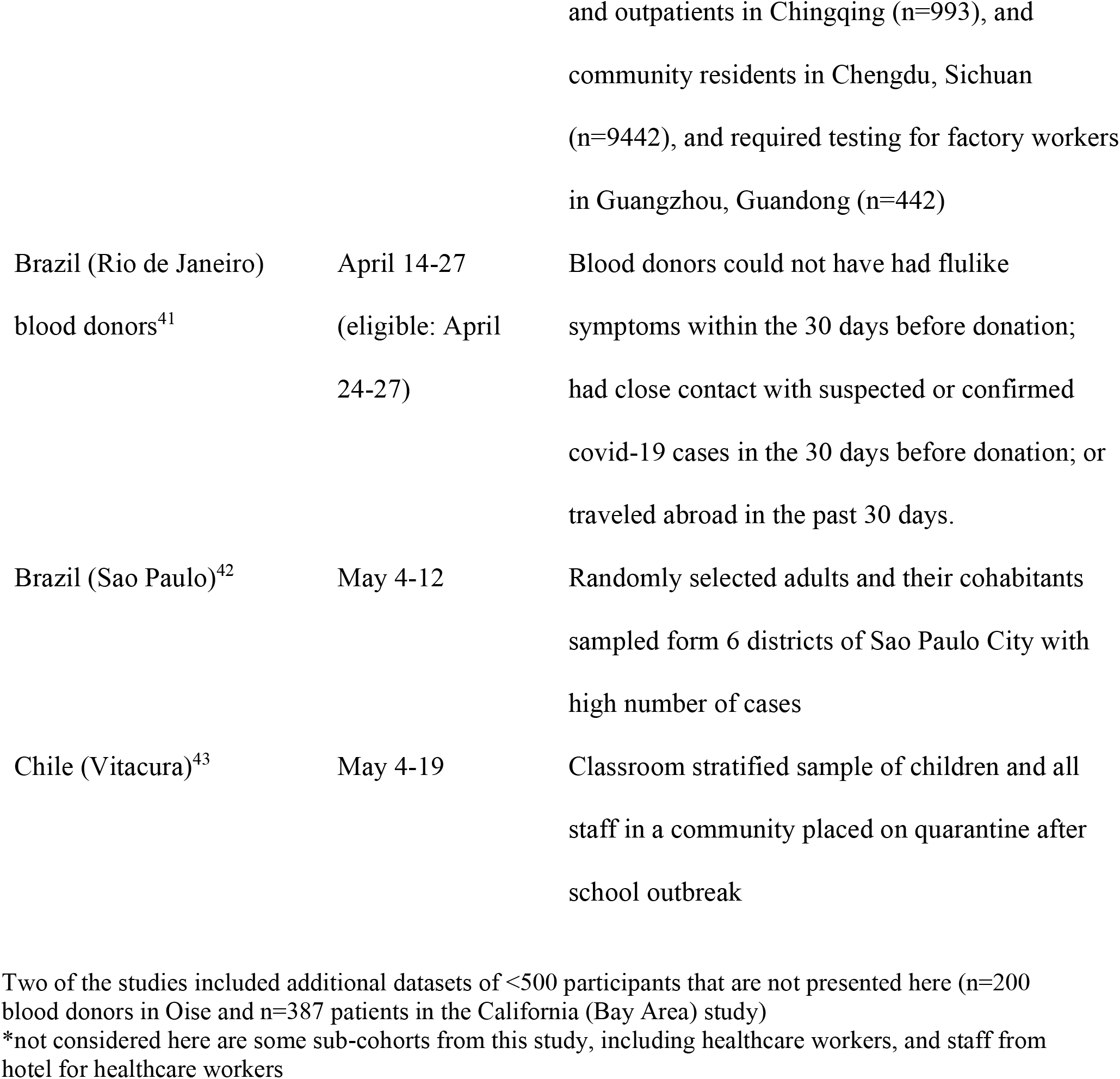
**Seroprevalence studies on COVID-19 published or depositing preprints as of July 11, 2020: dates, sampling and recruitment process**

**Table 2.** Sample size, types of antibodies, and population in relevant region.

| **Location** | **Sample size** | **Antibody** | **Population in region\*** | **Population**<br>**<70 years (%)** |
| --- | --- | --- | --- | --- |
| Iran (Guilan)<sup>8</sup> | 551 | IgG/IgM | 2354848 | 95 |
| Idaho (Boise)<sup>9</sup> | 4856 | IgG | 481587 (Ada county) | 92 |
| Switzerland (Geneva)<sup>10</sup> | 577 (4/20-27) | IgG | 500000 | 88 |
| Japan (Kobe)<sup>11</sup> | 1000 | IgG | 1518870 | 79 (Japan) |
| Denmark blood donors<sup>12</sup> | 20640 | IgG/IgM | 5771876 | 86 |
| France (Oise)<sup>13</sup> | 661 | IgG | 5978000 (Hauts-de-France) | 89 |
| China (Wuhan)<sup>14</sup> | 1401 | IgG/IgM | 11080000 | 93 (China) |
| Netherlands blood donors<sup>15</sup> | 7361 | IgG/IgM/IgA | 17097123 | 86 |
| Germany (Gangelt)<sup>16</sup> | 919 | IgG/IgA | 12597 | 86 |
| Brazil (Rio Grande do Sul)<sup>17</sup> | 4500 | IgG | 11377239 | 91 |
| Scotland blood donors<sup>18</sup> | 500 | IgG | 5400000 | 88 |
| California (Santa Clara)<sup>19</sup> | 3300 | IgG/IgM | 1928000 | 90 |
| Luxembourg<sup>20</sup> | 1807 | IgG/IgA** | 615729 | 90 |
| Germany (Frankfurt)<sup>21</sup> | 1000 | IgG | 2681000*** | 84 (Germany) |
| California (Los Angeles)<sup>22</sup> | 863 | IgG/IgM | 7892000 | 92 |
| New York<sup>23</sup> | 15101 | IgG | 19450000 | 90 |
| California (Bay Area)<sup>24</sup> | 1000 | IgG | 7753000 | 90 |
| Brazil (133 cities)<sup>25</sup> | 24995 | IgG/IgM | 74656499 | 94 (Brazil) |
| Croatia<sup>26</sup> | 1494 | IgG/IgM | 4076000 | 86 |
| New York (Brooklyn)<sup>27</sup> | 11092 | IgG | 2559903 | 91 |
| Switzerland (Zurich)<sup>28</sup> | 1644 patients (4/1-15) | IgG | 1520968 (canton Zurich)<br>1930525 (Zurich+Lucerne) | 88 |
|  | 1640 blood donors<br>(May) |  |  |  |
| Japan (Tokyo)<sup>29</sup> | 1071 | IgG | 13902077 | 79 (Japan) |
| Spain (Barcelona)<sup>30</sup> | 874 | IgG/IgM/IgA | 7566000 (Catalonia) | 86 |
| Italy (Apulia) blood donors<sup>31</sup> | 909 | IgG/IgM | 4029000 | 84 |
| China (Wuhan B)<sup>32</sup> | 1196 (4/4-8) | IgG/IgM | 11080000 | 93 (China) |
| California (San Francisco)<sup>33</sup> |  | IgG (also PCR testing) |  | 95 |
|  | 3953 |  | 5174 (census 022901) |  |
| Brazil (Espirito Santo)<sup>34</sup> | 4608 | IgG/IgM | 4018650 | 94 (Brazil) |
| USA (six states)<sup>35</sup> |  | Pan-Ig |  |  |
| WA Puget Sound | 3264 |  | 4273548 | 90 (WA) |
| UT | 1132 |  | 3282120 | 92 |
| NYC | 2482 |  | 9260870 | 89 |
| MO | 1882 |  | 6110800 | 88 |
| South FL | 1742 |  | 6345345 | 86 (FL) |
| CT | 1431 |  | 3562989 | 88 |
| Spain<sup>36</sup> | 61075 | IgG | 46940000 | 85 |
| Louisiana (Orleans and Jefferson Parish)<sup>37</sup> | 2640 | IgG | 825057 | 92 (LA) |
| Belgium<sup>38</sup> | 3391 (4/20-26) | IgG | 11589623 | 86 |
| France (Crepy-en-Valois)<sup>39</sup> | 1340 | IgG | 5978000 (Hauts-de-France) | 89 |
| China (several regions)<sup>40</sup> |  | IgG/IgM |  |  |
| Hubei (not Wuhan) | 979 |  | 48058000 | 93 (China) |
| Chongqing | 993 |  | 31243200 | 93 (China) |
| Sichuan | 9442 |  | 83750000 | 93 (China) |
| Guangdong | 1005 |  | 115210000 | 93 (China) |
| Brazil (Rio de Janeiro) | 669 (4/24-27) | IgG/IgM | 17264943 | 94 (Brazil) |
| blood donors<sup>41</sup> |  |  |  |  |
| Brazil (Sao Paulo)<sup>42</sup> | 517 | IgG/IgM | 298240 (6 districts) | 94 (Brazil) |
| Chile (Vitacura)<sup>43</sup> | 1244 | IgG/IgM | 85000 | 92 (Chile) |

\*The authors of some studies preferred to focus on age-restricted populations: 17-70 years old in the Denmark blood donor study (n=3800000), those 18-79 years old in the Luxembourg study (n=483000); those <70 years old in Netherlands blood donor study (n=13745768); those ≥18 years old in the New York state study (n=15280000); and those >19 years old in the UT population of the six states US study (n=2173082).
\*\*considered positive if both IgG and IgA were positive
\*\*\*participants were recruited from a large number of districts, but most districts had very few participants; here the population of the 9 districts with >1:10,000 sampling ratio is included (846/1000 participants came from these 9 districts).

**Table 3.** Prevalence of infection and estimated number of infected people.

| **Location** | **Seroprevalence (%)** |  | **Estimated infected** |
| --- | --- | --- | --- |
|  | **Crude** | **Adjusted (adjustments)** |  |
| Iran (Guilan)<sup>8</sup> | 22.0 | 33.0 (test, sampling) | 770000 |
| Idaho (Boise)<sup>9</sup> | 1.79 | ND | 8620 |
| Switzerland (Geneva)<sup>10</sup> | 10.6 | 10.9 (test, age, sex) | 54500 |
| Japan (Kobe)<sup>11</sup> | 3.3 | 2.7 (age, sex) | 40999 |
| Denmark blood donors<sup>12</sup> | 2.0 | 1.9 (test) | 109665 |
| France (Oise)<sup>13</sup> | 25.9 | ND | 1548000 |
| China (Wuhan)<sup>14</sup> | 10.0 | ND | 1108000 |
| Netherlands blood donors<sup>15</sup> | 2.7 | ND | 461622 |
| Germany (Gangelt)<sup>16</sup> | 15.0 | 20.0 (test, cluster, sym) | 2519 |
| Brazil (Rio Grande do Sul)<sup>17</sup> | 0.222 | 0.222 (sampling)* | 25283 |
| Scotland blood donors<sup>18</sup> | 1.2 | ND | 64800 |
| California (Santa Clara)<sup>19</sup> | 1.5 | 2.6 (test, sampling, cluster) | 51000 |
| Luxembourg<sup>20</sup> | 1.9 | 2.06 (age, sex, district) | 12684 |
| Germany (Frankfurt)<sup>21</sup> | 0.6 | ND | 16086 |
| California (Los Angeles)<sup>22</sup> | 4.06 | 4.65 (test, sex, race/ethnicity, income) | 367000 |
| New York<sup>23</sup> | 12.5 | 14.0 (test, sex, age race/ethnicity, region) | 2723000 |
| California (Bay Area)<sup>24</sup> | 0.4 (blood donors) | 0.1 (test/confirmation) | 7753 |
| Brazil (133 cities)<sup>25</sup> | 1.39 | 1.62 overall, varying from<br>0 to 25.0 across 133 cities<br>(test, design) | 1209435** |
| Croatia<sup>26</sup> | 1.27*** | ND | 51765 |
| New York (Brooklyn)<sup>27</sup> | 47.0 | ND | 1203154 |
| Switzerland (Zurich)<sup>28</sup> | unclear | 1.3 in patients in April 1-15 and 1.6 in blood<br>donors in May<br>(multivariate Gaussian<br>conditioning) | 19773 (Zurich)<br>30888<br>(Zurich+Lucerne) |
| Japan (Tokyo)<sup>29</sup> | 3.83 | ND | 532450 |
| Spain (Barcelona)<sup>30</sup> | 14.3 | ND | 1081938 |
| Italy (Apulia) blood<br>donors<sup>31</sup> | 0.99 | ND | 39887 |
| China (Wuhan B)<sup>32</sup> | 8.36 (3.53 for entire<br>period) | ND (2.80 (age, gender,<br>test) for entire period) | 926288 |
| California (San Francisco)<sup>33</sup> | 4.3 in the census track | 6.1 (age, sex,<br>race/ethnicity, test) | 316 |
| Brazil (Espírito Santo)<sup>34</sup> | 2.1 | ND | 84391 |
| USA (six states)<sup>35</sup> |  | (age, sex, test) |  |
| WA Puget Sound | 1.3 | 1.13 | 48291 |
| UT | 2.4 | 2.18 | 71550 |
| NYC | 5.7 | 6.93 | 641778 |
| MO | 2.9 | 2.65 | 161936 |
| South FL | 2.2 | 1.85 | 117389 |
| CT | 4.9 | 4.94 | 176012 |
| Spain<sup>36</sup> | ND | 5.0**** (sampling, age, sex, income) | 2347000 |
| Louisiana (Orleans and Jefferson Parish)<sup>37</sup> | 6.9 (IgG or PCR) | 6.9 for IgG (census weighting, demographics) | 56578 |
| Belgium<sup>38</sup> | 5.7 | 6.0 (sampling, age, sex, province) | 695377 |
| France (Crepy-en-Valois)<sup>39</sup> | 10.4 | ND | 620105 |
| China (several regions)<sup>40</sup> |  |  |  |
| Hubei (not Wuhan) | 3.6 | ND | 1718110 |
| Chongqing | 3.8 | ND | 11956109 |
| Sichuan | 0.6 | ND | 487847 |
| Guangdong | 2.2 | ND | 2522010 |
| Brazil (Rio de Janeiro) blood donors<sup>41</sup> | 6.0 | 4.7 (age, sex, test) | 811452 |
| Brazil (Sao Paulo)<sup>42</sup> | 5.2 | 4.7 (sampling design) | 14017 |
| Chile (Vitacura)<sup>43</sup> | 11.2 | ND | 9500 |

Among studies with multiple consecutive time points where seroprevalence was evaluated, the seroprevalence estimate was the highest in the most recent time interval with the exception of Switzerland (Geneva) where the highest value was seen two weeks before the last time interval, Switzerland (Zurich) where the highest was seen in April 1-15 for patients at the university hospital and in May for blood donors, and the China (Wuhan B) study where the highest value was seen about 3 weeks before the last time interval.
\*an estimate is also provided adjusting for test performance, but the assumed specificity of 99.0% seems inappropriate, since as part of the validation process the authors also found that several of the test-positive individuals had household members who were also infected, thus the estimated specificity was deemed to be at least 99.95%
\*\*the authors calculate 760000 infected in the 90 cities that had 200-250 samples tested, but many of the other 43 cities with <200 samples may be equally or even better represented, since they tended to be smaller than the 90 (mean population 356213 versus 659326)
\*\*\*1.20% in workers in Split without mobility restrictions, 3.37% in workers in Knin without mobility restrictions, 1.57% for all workers without mobility restrictions; Split and Knin tended to have somewhat higher death rates than nation-wide Croatia, but residence of workers is not given, so the entire population of the country is used in the calculations
\*\*\*\* 5.0% by point of care test, 4.6% with immunoassay, 3.7% by having both tests positive, 6.2% by having at least one test positive
test: test performance; ND: no data available

**Table 4.**
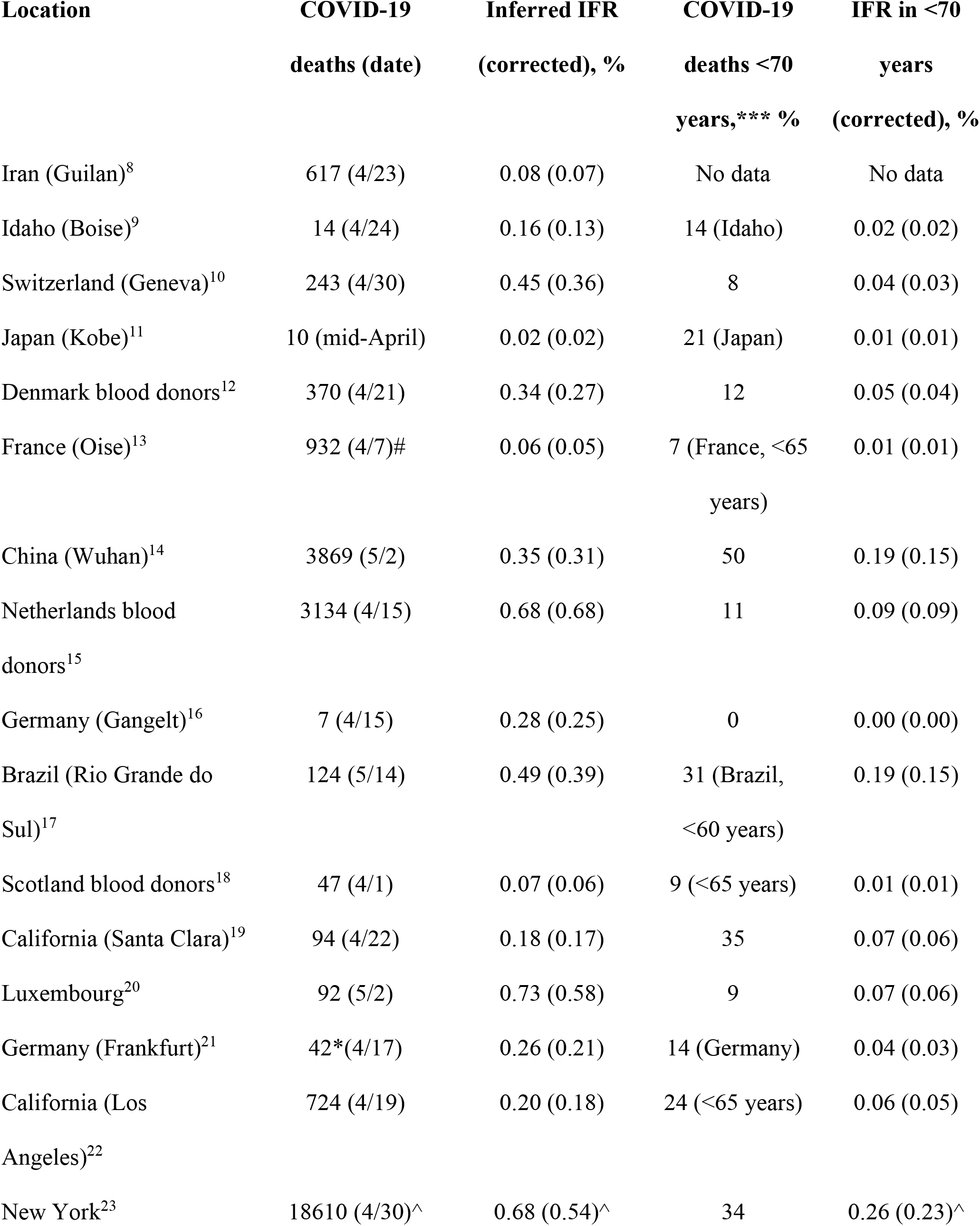
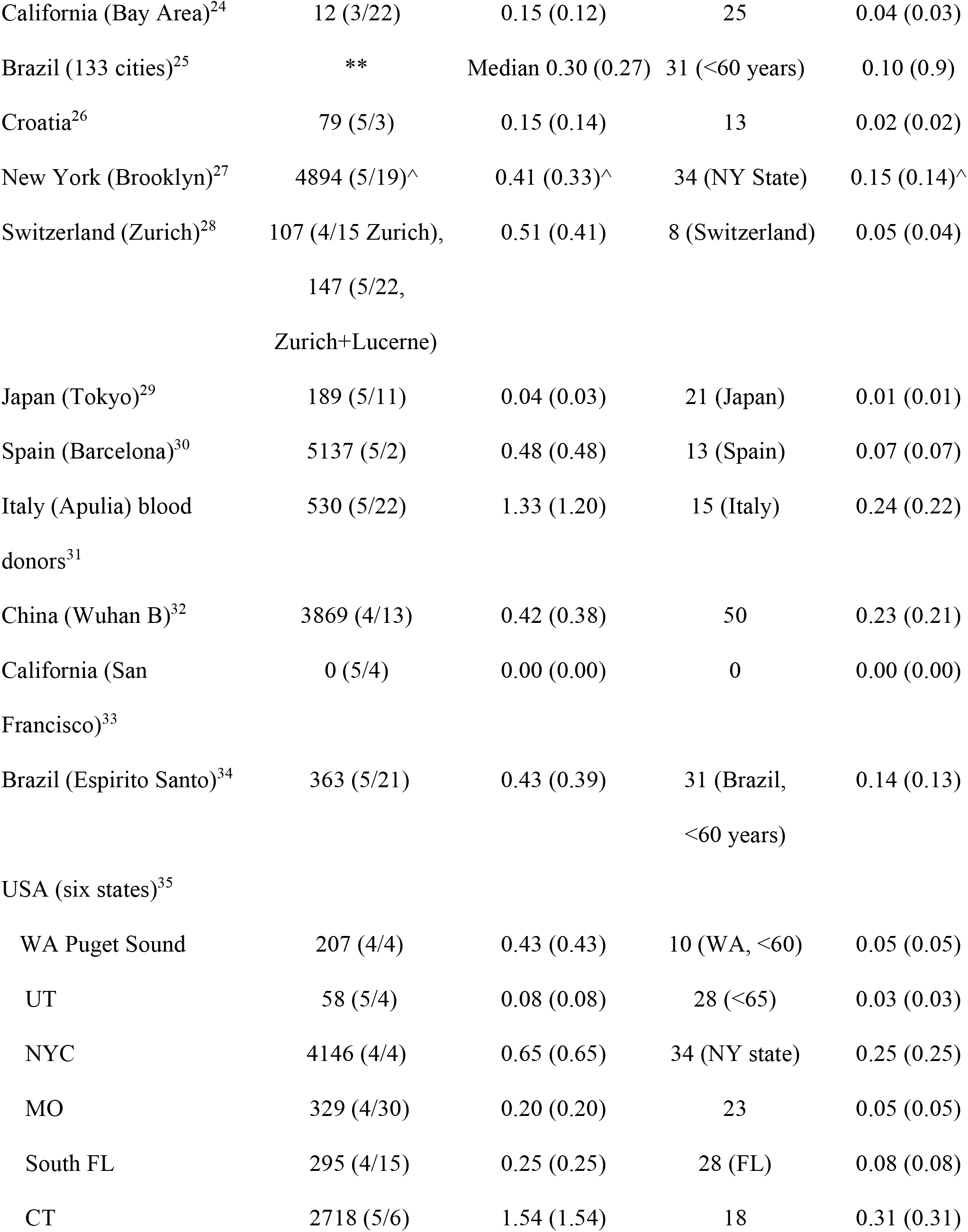
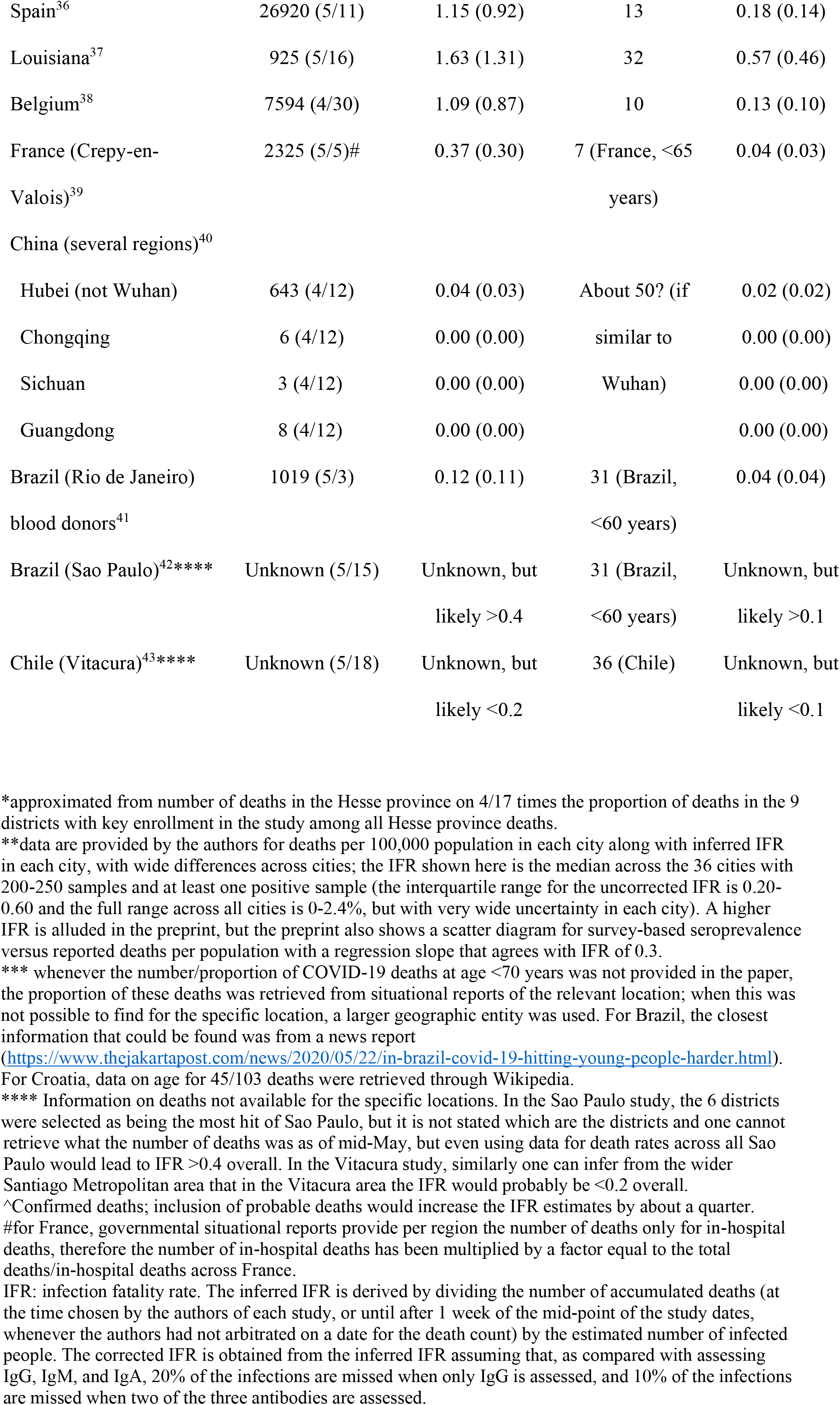
Inferred infection fatality rates.

**Table 5.** **Additional seroprevalence data from nation-wide studies that have been announced to the press and/or in preliminary reports, but are not presented yet as full articles**. Only countries not represented in Tables 1-4 are considered

| Country | Sample size<br>(antibody) | Date | Seroprevalence<br>(%) | Population | Deaths (date) | IFR (corrected) |
| --- | --- | --- | --- | --- | --- | --- |
| United Kingdom<sup>44</sup> | 885 (IgG) | April 26-May 24<sup>^</sup> | 6.78 | 67897000 | 34636 (5/17) | 0.75 (0.60) |
| Finland<sup>45</sup> | 674 (IgG) | April 20-26<sup>^</sup> | 2.52 | 5541000 | 211 (4/30) | 0.15 (0.12) |
| Sweden<sup>46</sup> | 1200 (IgG) | May 18-24 | 6.3 | 10101000 | 4501 (5/28) | 0.71 (0.57) |
| Czechia<sup>47</sup> | 26549 (IgG) | April 23-May 1 | 0.40 | 10710000 | 252 (5/4) | 0.59 (0.47) |
| Israel<sup>48</sup> | 1709 (IgG?) | May 2020 | 2-3 | 9198000 | 299 (6/10) | 0.13 (0.10)* |
| India<sup>49</sup> | 26400 (IgG?) | May 2020 | 0.73** | 1380382000 | 8107 (6/10) | 0.08 (0.06) |
| Slovenia<sup>50</sup> | 1368 (IgG?) | April 2020 | 3.1 | 2079000 | 92 (5/1) | 0.14 (0.11) |

<sup>^</sup> slightly lower seroprevalence recorded in subsequent weeks
\*Assuming seroprevalence 2.5%. Another ongoing study in Beni Brak (population n=198900, 0 deaths as of mid-June) has shown seroprevalence of 1.4% among the first 2933 samples. A much larger seroprevalence study based on HMOs has also been launched.
\*\*based on data from part of the sample (65 of the 83 districts)

At least seven studies found some statistically significant, modest differences in seroprevalence rates across some age groups (Oise: decreased seroprevalence in age 0-14, increased in age 15-17; Geneva: decreased seroprevalence in age >50; Netherlands: increased seroprevalence in age 18-30; New York state: decreased seroprevalence in age >55; Brooklyn: decreased seroprevalence in age 0-5, increased in age 16-20; Tokyo: increased seroprevalence in age 18-34, Spain: decreased seroprevalence in age 0-10, Belgium: higher seroprevalence in age >90). The patterns are not strong enough to suggest major differences in extrapolating across age groups, although higher values in adolescents and young adults and lower values in children cannot be excluded.

As shown in Table 1, these studies varied substantially in sampling and recruitment designs. The main issue is whether they can offer a representative picture of the population in the region where they are performed. A generic problem is that vulnerable people who are at high risk of infection and/or death may be more difficult to recruit in survey-type studies. COVID-19 infection seems to be particularly widespread and/or lethal in nursing homes, among homeless people, in prisons, and in disadvantaged minorities. Most of these populations are very difficult, or even impossible to reach and sample from and they are probably under-represented to various degrees (or even entirely missed) in surveys. This would result in an underestimation of seroprevalence and thus overestimation of IFR. Eleven of the 36 studies that are available as full papers (Iran,^8^ Geneva,^10^ Gangelt,^16^ Rio Grande do Sul,^17^ Luxembourg,^20^ Los Angeles county,^22^ three Brazil studies,^25,34,42^, Spain,^36^ and Louisiana^37^) explicitly aimed for random sampling from the general population. In principle, this is a stronger design. However, even with such designs, people who cannot be reached (e.g. by e-mail or phone or even visiting them at a house location) will not be recruited, and these vulnerable populations are likely to be missed. Moreover, 5 of these 11 studies^8,10,16,42,37^ focused on studying geographical locations that had extreme numbers of deaths, higher than other locations in the same city or country, and this would tend to select eventually for higher IFR on average.

Seven studies assessed blood donors in Denmark,^12^ Netherlands,^15^ Scotland,^18^ the Bay Area in California,^24^ Zurich/Lucerne,^28^ Apulia^31^ and Rio De Janeiro.^41^ By definition these studies include people in good health and without symptoms, at least recently, and therefore may markedly underestimate COVID-19 seroprevalence in the general population. A small set of 200 blood donors in Oise, France^13^ showed 3% seroprevalence, while pupils, siblings, parents, teachings and staff at a high school with a cluster of cases in the same area had 25.9% seroprevalence; true population seroprevalence may be between these two values.

For the other studies, healthy volunteer bias may lead to underestimating seroprevalence and this is likely to have been the case in at least one case (the Santa Clara study)^19^ where wealthy healthy people were rapidly interested to be recruited when the recruiting Facebook ad was released. The design of the study anticipated correction with adjustment of the sampling weights by zip code, gender, and ethnicity, but it is likely that healthy volunteer bias may still have led to some underestimation of seroprevalence. Conversely, attracting individuals who might have been concerned of having been infected (e.g. because they had symptoms) may lead to overestimation of seroprevalence in surveys. Finally studies of employees, grocery store clients, or patient cohorts (e.g. hospitalized for other reasons, or coming to the emergency room, or studies using residual lab samples) may have sampling bias with unpredictable direction.

As shown in Table 2, all studies have tested for IgG antibodies, but only about half have also assessed IgM, 4 have assessed IgA. Only three studies assessed all three types of antibodies and one more used a pan-Ig antibody. Studies typically considered the results to be “positive” if any tested antibody type was positive, but one study (Luxembourg) that considered the results to be “positive” only if both IgG and IgA were detected. The ratio of people sampled versus the total population of the region was better than 1:1000 in 11 studies (Idaho,^9^ Denmark blood donors,^12^ Gangelt,^16^ Santa Clara,^19^ Luxembourg,^20^ Brooklyn,^27^ Zurich,^28^ San Francisco,^33^ Espirito Santo,^34^ Spain,^36^ and Vitacura^43^).

### Seroprevalence estimates

As shown in Table 3, prevalence ranged from as little as 0.222% to as high as 47%. Studies varied a lot on whether they tried or not to adjust their estimates for test performance, sampling (striving to get closer to a more representative sample), and clustering effects (e.g. when including same household members) as well as other factors. The adjusted seroprevalence occasionally differed substantially from the crude, unadjusted value. In principle adjusted values are likely to be closer to the true estimate, but the exercise shows that each study alone may have some unavoidable uncertainty and fluctuation, depending on the analytical choices preferred. In studies that sampled people from multiple locations, large between-location heterogeneity could be seen (e.g. 0-25% across 133 Brazilian cities).^25^

### Inferred IFR

Inferred IFR estimates varied a lot, from 0.00% to 1.63%. Corrected values also varied extensively, from 0.00% to 1.31%. For 10 locations, more than one IFR estimate was available and thus IFR from different studies evaluating the same location could be compared. As shown in figure 1, the IFR estimates tended to be more homogeneous within each location, while they differed a lot across locations. The sample size-weighted summary was used to generate a single estimate to represent each location. Data were available for 32 different locations. The median IFR across all 32 locations was 0.27% (0.24% using the corrected values). Most data came from locations with high death tolls and 23 of the 32 locations had a population mortality rate (deaths per million population) higher than the global average (73 deaths per million population as of July 12) (Figure 2). The uncorrected IFR estimates had a range of 0.01-0.16% (median 0.13%) across the 9 locations with population mortality rate below the global average, 0.07-0.73% (median 0.27%) across the 15 locations with population mortality rate above the global average but below 500 deaths per million population, and 0.59-1.63% (median 1.12%) across the 8 extreme hotbed locations with over 500 deaths per million population. The corrected IFR estimates had medians of 0.10%, 0.25%, and 0.90%, respectively, for the three groups of locations.

**Figure 1.**
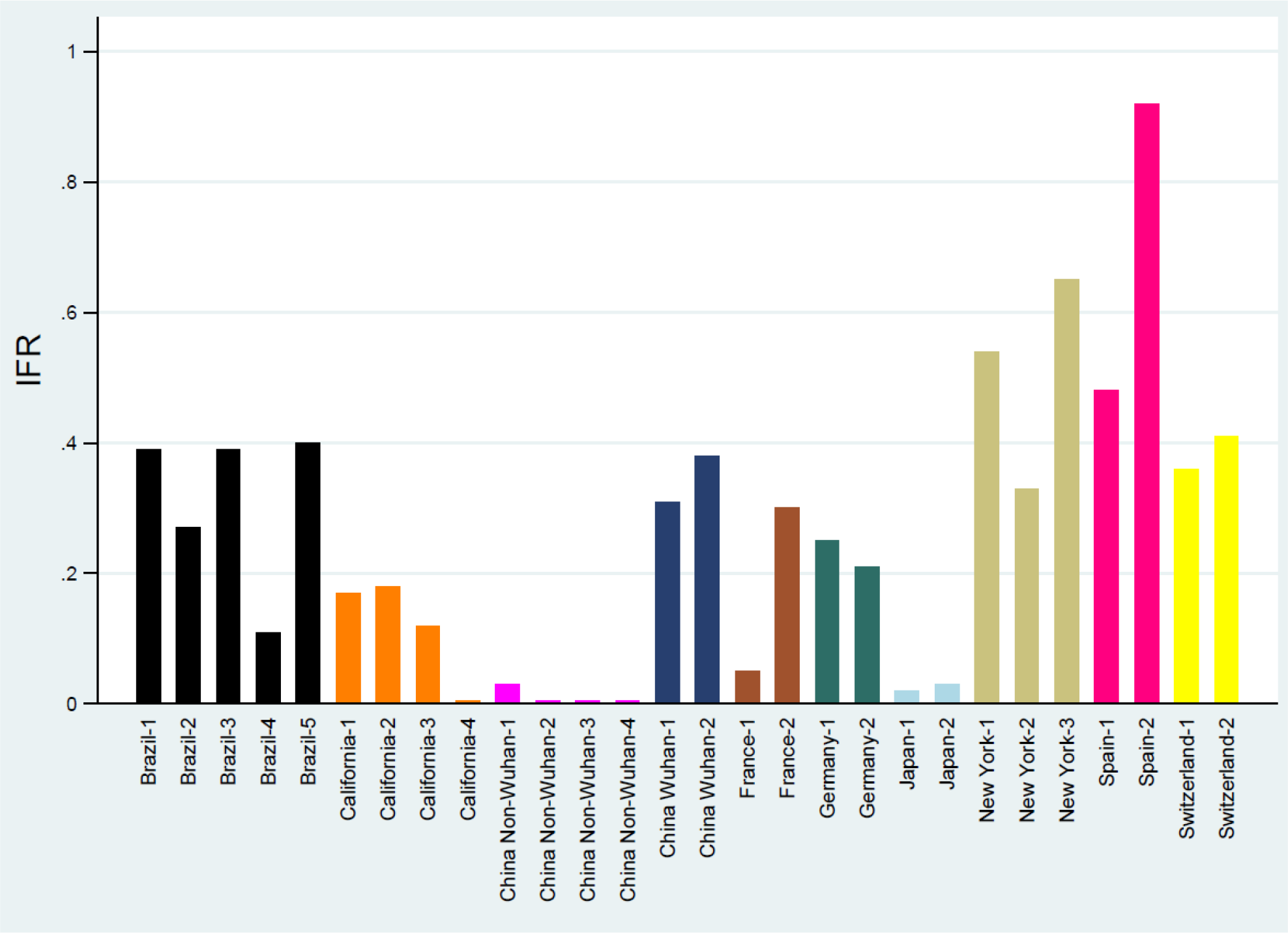
Locations that had two or more IFR estimates from different studies. Locations are defined at the level of countries, except for the USA where they are defined at the level of states and China is separated into Wuhan and non-Wuhan areas. Corrected IFR estimates are shown. Within the same location, IFR estimates tend to have only modest differences, even though it is possible that different areas within the same location may also have genuinely different IFR. France is one exception where differences are large, but both estimates come from population studies of outbreaks from schools and thus may not provide good estimates of population seroprevalence and may lead to underestimated IFR.

**Figure 2.**
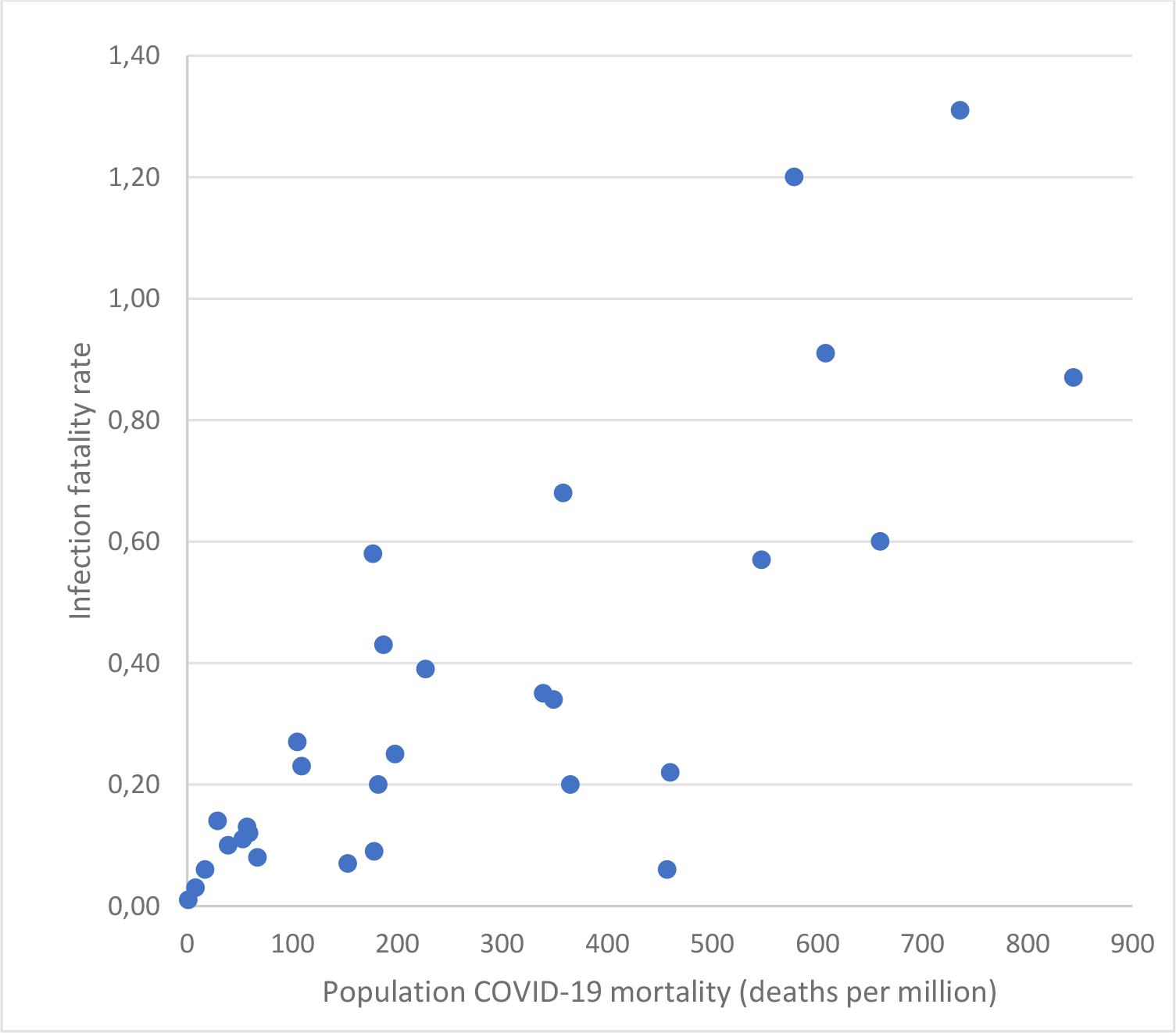
Scatterplot of corrected IFR estimates (%) in each location plotted against the COVID-19 mortality rate as of July 12, 2020 in that location (in deaths per million population). Locations are defined at the level of countries, except for the USA where they are defined at the level of states and China is separated into Wuhan and non-Wuhan areas. When several IFR estimates are available from multiple studies for a location, the sample size-weighted mean has been used. Not shown are two locations with >1000 deaths per million population, both of which have high IFR (New York and Connecticut).

The proportion of COVID-19 deaths that occurred in people <70 years old varied substantially across locations. All deaths in Gangelt were in elderly people while in Wuhan half the deaths occurred in people <70 years old and the proportion might have been higher in Iran, but no data could be retrieved for this country. When limited to people <70 years old, IFR ranged from 0.00% to 0.57% with median of 0.05% (corrected, 0.00-0.46% with median of 0.04%). IFR estimates in people <70 years old were lower than 0.1% in all but 7 locations that were hard-hit hotbeds (Belgium, Wuhan, Italy, Spain, Connecticut, Louisiana, New York).

## DISCUSSION

IFR is not a fixed physical constant and it can vary substantially across locations, depending on the population structure, the case-mix of infected and deceased individuals and other, local factors. Inferred IFR values based on emerging seroprevalence studies typically show a much lower fatality than initially speculated in the earlier days of the pandemic.

The studies analyzed here represent 50 different estimates of IFR, but they are not fully representative of all countries and locations around the world. Most of them come from locations with overall COVID-19 mortality rates exceeding the global average (73 deaths per million people as of July 12). The median inferred IFR in locations with COVID-19 mortality rate below the global average is low (0.13%, corrected 0.10%). For hotbed countries with COVID-19 mortality rates above the global average but lower than 500 deaths per million, the median IFR is still not that high (median 0.27%, corrected 0.25%). Very high IFR estimates have been documented practically in locations that had devastating experiences with COVID-19. Such epicenters are unusual across the globe, but they are overrepresented in the 50 seroprevalence estimates available for this analysis. Therefore, if one could sample equally from all countries and locations around the globe, the median IFR might be even lower than the one observed in the current analysis.

Several studies in hard-hit European countries inferred modestly high IFR estimates for the overall population, but the IFR was still low in people <70 years old. Some of these studies were on blood donors and may have underestimated seroprevalence and overestimated IFR. One study in Germany aimed to test the entire population of a city and thus selection bias is minimal: Gangelt^16^ represents a situation with a superspreader event (in a local carnival) and 7 deaths were recorded, all of them in very elderly individuals (average age 81, sd 3.5). COVID-19 has a very steep age gradient of death risk.^51^ It is expected therefore that in locations where the infection finds its way into killing predominantly elderly citizens, the overall, age-unadjusted IFR would be higher. However, IFR would still be very low in people <70 in these locations, e.g. in Gangelt IFR is 0.00% in non-elderly people. Similarly, in Switzerland, 69% of deaths occurred in people >80 years old^51^ and this explains the relatively high overall IFR in Geneva and Zurich. Similar to Germany, very few deaths in Switzerland have been recorded in non-elderly people, e.g. only 2.5% have occurred in people <60 years old and IFR in that age-group would be ∼0.01%. The majority of deaths in most of the hard hit European countries have happened in nursing homes^52^ and a large proportion of deaths also in the US^53^ also follow this pattern. Moreover, many nursing home deaths have no laboratory confirmation and thus should be seen with extra caution in terms of the causal impact of SARS-CoV-2.

Locations with high burdens of nursing home deaths may have high IFR estimates, but the IFR would still be very low among non-elderly, non-debilitated people. The average length of stay in a nursing home is slightly more than 2 years and people who die in nursing homes die in a median of 5 months^54^ so many COVID-19 nursing home deaths may have happened in people with life expectancy of only a few months. This needs to be verified in careful assessments of COVID-19 outbreaks in nursing homes with detailed risk profiling of fatalities. If COVID-19 happened in patients with very limited life expectancy, this pattern may even create a dent of less than expected mortality in the next 3-6 months after the coronavirus excess mortality wave. As of July 12 (week 28), preliminary Euromonitor data^55^ indeed already show a substantial dent below baseline mortality in France, and a dent below baseline mortality is seen also for the aggregate European data.

Within China, the much higher IFR estimates in Wuhan versus other areas may reflect the wide spread of the infection to hospital personnel and the substantial contribution of nosocomial infections to a higher death toll in Wuhan;^56^ plus unfamiliarity with how to deal with the infection in the first location where COVID-19 arose. Massive deaths of elderly individuals in nursing homes, nosocomial infections, and overwhelmed hospitals may also explain the very high fatality in specific locations in Italy^57^ and in New York and neighboring states. Seroprevalence studies in health care workers and administrative hospital staff in Lombardy^58^ found 8% seroprevalence in Milan hospitals and 35-43% in Bergamo hospitals, supporting the scenario for widespread nosocomial infections among vulnerable patients. The high IFR values in New York metropolitan area and neighboring states are also not surprising, given the vast death toll witnessed. A very unfortunate decision of the several state governors was to have COVID-19 patients sent to nursing homes. Moreover, some hospitals in New York City hotspots reached maximum capacity and perhaps could not offer optimal care. Use of unnecessarily aggressive management (e.g. mechanical ventilation) and hydroxychloroquine may also have contributed to worse outcomes. Furthermore, New York City has an extremely busy, congested public transport system that may have exposed large segments of the population to high infectious load in close contact transmission and, thus, perhaps more severe disease. A more aggressive viral clade has also been speculated, but this needs further verification.^59^

IFR may reach very high levels among disadvantaged populations and settings that have the worst combination of factors predisposing to higher fatalities. Importantly, such hotspot locations are rather uncommon exceptions in the global landscape. Moreover, even in these locations, the IFR for non-elderly individuals without predisposing conditions may remain very low. E.g. in New York City only 0.65% of all deaths happened in people <65 years without major underlying conditions.^51^ Thus the IFR even in New York City would probably be lower than 0.01% in these people.

Studies with extremely low inferred IFR are also worthwhile discussing. Possible overestimation of seroprevalence and undercounting of deaths need to be considered. E.g., for Kobe, the authors of the study^11^ raise the question whether COVID-19 deaths have been undercounted in Japan. Both undercounting and overcounting of COVID-19 deaths may be a caveat in different locations and this is difficult to settle in the absence of very careful scrutiny of medical records and autopsies. The Tokyo data,^29^ nevertheless, also show similarly very low IFR. Moreover, evaluation of all-cause mortality in Japan has shown no excess deaths during the pandemic, consistent with the possibility that somehow the Japanese population was spared. Very low IFRs seem common in Asian countries, including China (excluding Wuhan), Iran, Israel and India. Former immunity from exposure to other coronaviruses, genetic differences, hygienic etiquette, lower infectious load, and other unknown factors may be speculated. IFR seems to be very low also in Singapore where extensive PCR testing was carried out. As of July 12, 2020, in Singapore there were only 26 deaths among 46,283 cases, suggesting an upper bound of 0.06% for IFR, even if no cases had been missed.

Some surveys have also been designed to assess seroprevalence repeatedly spacing out measurements in the same population over time. A typical pattern that seems to emerge is that seroprevalence may increase several fold within a few weeks, but plateau or even decline may follow.^10,28^ A more prominent decline of seropositivity was seen in a study in Wuhan.^32^ Genuine decrease may be difficult to differentiate from random variation. However, some preliminary data^60,61^ suggest that decrease in antibody titers may be fast. Decrease in seropositivity over time means that the numbers of infected people may be underestimated and IFR overestimated.

The only data from a low-income country among the 23 studies examined here come from Iran^8^ and India^49^ and the IFR estimates appears to be very low. Iran has a young population with only slightly over 1% of the age pyramid at age >80 and India’s population is even younger. Similar considerations apply to almost every less developed country around the world. Given the very sharp age gradient and the sparing of children and young adults from death by COVID-19, one may expect IFR to be fairly low in the less developed countries. However, it remains to be seen whether comorbidities, poverty and frailty (e.g. malnutrition) and congested urban living circumstances may have adverse impact on risk and thus increase IFR also in these countries.

One should caution that the extent of validation of the antibody assays against positive and negative controls differs across studies. Specificity has typically exceeded 99.0%, which is reassuring. However, for very low prevalence rates, even 99% specificity may be problematic. Sensitivity also varies from 60-100% in different validation exercises and for different tests, but typically it is closer to the upper than the lower bound. One caveat about sensitivity is that typically the positive controls are patients who had symptoms and thus were tested and found to be PCR-positive. However, it is possible that symptomatic patients may be more likely to develop antibodies than patients who are asymptomatic or have minimal symptoms and thus had not sought PCR testing.^61-65^ For example, one study found that 40% of asymptomatic patients became seronegative within 8 weeks.^61^ Since the seroprevalence studies specifically try to unearth these asymptomatic/mildly symptomatic missed infections, a lower sensitivity for these mild infections could translate to substantial underestimates of the number of infected people and substantial overestimate of the inferred IFR.

The corrected IFR estimates are trying to account for undercounting of infected people when not all 3 antibodies (IgG, IgM, and IgA) are assessed.^7^ However, the magnitude of the correction is uncertain and may also vary in different circumstances. Moreover, it is possible that an unknown proportion of people may have handled the virus using immune mechanisms (mucosal, innate, cellular) that did not generate any serum antibodies.^66-69^ This may lead to substantial underestimation of the frequency of infection and respective overestimation of the IFR. One study has found indeed that mild SARS-CoV-2 infections may lead to nasal release of IgA, without serum antibody response.^68^ Another study has found that 6 of 8 interfamilial contacts of index cases remained seronegative despite developing symptoms and 6 of 8 developed persisting T cell responses^69^ and the important role of cellular immune responses even in seronegative patients has been documented also by other investigators.^70^

An interesting observation is that even under congested circumstances, like cruise ships, aircraft carriers or homeless shelter, the proportion of people detected positive typically does not get to exceed 20-45%.^71,72^ Similarly, at a wider population level, values ∼47% are the maximum values documented to-date and most values are much lower, yet epidemic waves seem to wane. It has been suggested^73,74^ that differences in host susceptibility and behavior can result in herd immunity at much lower prevalence of infection in the population than originally expected. COVID-19 spreads by infecting certain groups more than others because some people have much higher likelihood of exposure. People most likely to be exposed also tend to be those most likely to spread for the same reasons that put them at high exposure risk. In the absence of random mixing of people, the epidemic wave may be extinguished even with relatively low proportions of people becoming infected. Seasonality may also play a role in the dissipation of the epidemic wave. It has also been observed that many people have CD4 cellular responses to SARS-CoV-2 even without being exposed to this virus, perhaps due to prior exposure to other coronaviruses.^75^ It is unknown whether this proportion varies in different populations around the world and whether this immunity may contribute to SARS-CoV-2 epidemic waves waning without infecting a large share of the population.

A major limitation of the current analysis is that the calculations presented in this paper include several studies that have not yet been fully peer-reviewed. Moreover, there are several studies that are still ongoing. New emerging data may offer more insights and updated estimates. Given that the large majority of studies have been done in locations that were hard hit from COVID-19, it would be useful to do more studies in less hit locations, so as to have a more balanced global perspective.

A comparison of COVID-19 to influenza is often attempted, but many are confused by this comparison unless placed in context. Based on the IFR estimates obtained here, COVID-19 may have infected as of July 12 approximately 300 million people (or more), far more than the ∼13 million PCR-documented cases. The global COVID-19 death toll is still evolving, but it is still not much dissimilar to a typical death toll from seasonal influenza (290,000-650,000),^76^ while “bad” influenza years (e.g. 1957-9 and 1968-70) have been associated with 1-4 million deaths.^77^ Notably, influenza devastates low-income countries, but is more tolerant of wealthy nations, probably because of the availability and wider use of vaccination in these countries.^58^ Conversely, in the absence of vaccine and with a clear preference for elderly debilitated individuals, COVID-19 may have an inverse death toll profile, with more deaths in wealthy nations than in low-income countries. However, even in the wealthy nations, COVID-19 seems to affect predominantly the frail, the disadvantaged, and the marginalized – as shown by high rates of infectious burden in nursing homes, homeless shelters, prisons, meat processing plants, and the strong racial/ethnic inequalities against minorities in terms of the cumulative death risk.^78,79^

While COVID-19 is a formidable threat, the fact that its IFR is typically much lower than originally feared, is a welcome piece of evidence. The median IFR found in this analysis is very similar to the estimate recently adopted by CDC for planning purposes.^80^ The fact that IFR can vary substantially also based on case-mix and settings involved also creates additional ground for evidence-based, more precise management strategies. Decision-makers can use measures that will try to avert having this lethal virus infect people and settings who are at high risk of severe outcomes. These measures may be possible to be more precise and tailored to specific high-risk individuals and settings than blind lockdown of the entire society. Of course, uncertainty remains about the future evolution of the pandemic, e.g. the presence and height of subsequent waves.^81^ However, it is helpful to know that SARS-CoV-2 has relatively modest IFR overall and that possibly IFR can be made even lower with appropriate, precise non-pharmacological choices.

## Data Availability

All data are included in the manuscript

